# Does mother’s pregnancy intention affect child healthcare and growth in India? Evidence from a longitudinal survey

**DOI:** 10.1101/2020.08.12.20149732

**Authors:** Poulomi Chowdhury, Mausam Kumar Garg, Md Illias Kanchan Sk

## Abstract

**Objective:** To assess the effect of unintended births on children’s health care and nutritional status.

**Design:** The study uses cross-sectional prospective design.

**Setting:** The nationally representative Indian Human Development Survey (IHDS) data of two rounds (i.e. 2004-5, 2011-12) was used in this study. The women data was used to draw representative sample of 7,166, and out of them 3,905 belong to under 5 years of age group.

**Statistical Analyses:** Secondary analysis, using bivariate and multivariate linear and logistic models was conducted using both rounds of IHDS data. We categorized birth as an unintended birth if mother did not want to have addition child at IHDS-I but gave birth during inter-survey. Furthermore, all births exceeding to the desired number of children reported by mothers in the IHDS-II were also included in unintended births and all other births were considered as intended births. Multivariate logistic regression models were applied to analyse the effect of unintended and intended births on child-immunization and exclusive breastfeeding. While multivariate linear regression models were used to assess the effect of childbearing intention on child nutritional status.

**Results:** The study shows that by controlling other factors, unintended births were less likely to be exclusively breastfed (OR 0.885, p<0.05) and receive full immunization (OR 0.830, p<0.001). Moreover, children’s poor nutrition was more prevalent among unintended births as they were more likely to be stunting and underweight.

**Conclusions:** The findings of the study underscore the importance of investments in family planning to reduce the unintended births to improve children’s health and growth.

**Strengths and limitations of the study:**

- The findings of this study are from nationally representative sample of 3,905 belong to under 5 years of age group.
- Other child development indicators namely cognitive skills and academic performance are not included in this study.
- The seven years of duration between the survey period could have changed women’s childbearing intention.

## INTRODUCTION

Unintended pregnancies are those pregnancies which occur at the time of conception, it is either mistimed or unwanted. It marks the serious socio-economic and health consequences for women and their families (1). It is observed that out of 213 million pregnancies worldwide 40% of them were unintended (2). Further, half of the unplanned births ended up in induced abortion (2), which is not safe in many countries that have the restricted abortion laws. Due to unsafe abortion women tend to face serious complication like haemorrhage (heavy bleeding), incomplete abortion (failure to remove all of the pregnancy tissue from uterus), infection, urine perforation, damage genital tract and internal organs (3). Subsequently, unintended pregnancies often result in unplanned births and miscarriages. Therefore, preventing unintended pregnancies is important in achieving the UN millennium development goal i.e. improving the maternal health (goal 5).

It is observed from various studies that children born from unintended pregnancies suffer from various social and health problems. These children receive inadequate prenatal care (4, 5) and immunization. They do not even receive proper exclusive breast-feeding, or refrain from doing so (6) resulting poor physical health that than those who are intended (4). Children from unintended pregnancies are associated with 1.41 times odds of having low birth weight as well as high risk of prematurity (7). Likewise, they have less close mother child relationship and poorer educational and behavioural outcomes (8).

India is the second most populous country in the world with a population of 1.3 billion. The National Family and Health Survey (NFHS-3) conducted in 2015-16 report reflects that around 35.8% of children under 5 years are underweight and 58.6% are anaemic. The percentage of children underweight varies differently across the states, it is highest in Bihar (43.9%), Jharkhand (47.8%) and Madhya Pradesh (42.8%) and lowest in Mizoram (12%), Manipur (13.8%) and Sikkim (14.2%). The prevailing poor health status of children in the country may become more cumbersome when unintended births come into the picture. Following the 21% unintended pregnancy as per the NFHS-3 (2005-06) it would be very important to study the its associated effect on children’s health. Recently, based on retrospective and prospective data of India, it is observed that unintended pregnancies have adverse effect on childhood vaccination and stunting (9, 10). Further, there are no current studies which have worked on the national level considering all the states of India and most research examining correlates of the intention status of women’s pregnancies in India has been based on data collected from cross-sectional surveys and using retrospective fertility intentions. Moreover, these studies do not provide the current scenario of unintended births. Therefore, this study analyses the effect of unintended births (measured using prospective approach) on child healthcare and growth in India using nationally representative prospective cross-sectional data.

## METHODS

### Study design

The quantitative approach has been used for this study. It focuses on unintended births and associated child health outcomes. Further, to emphasis on unintended births at national level, the prospective cross-sectional data is the suitable option.

### Data source

The present study utilized data from two rounds of the Indian Human Development Survey (IHDS), conducted during 2005-06 and 2011-12. IHDS covered 41,554 households in round 1 and 42,152 households in round 2. IHDS is nationally representative survey which provides data on different topics such as health, education, employment, economic status, marriage, fertility, gender relations, and social capital. The data of completed short reading, writing and arithmetic tests of children aged 8-11 is also available (11).

For this study, the women data file of two rounds of IHDS was linked together. A total number of women accounted for 25,479 who were available in both the rounds were considered for the analysis. Then, children file was created by combining ever-married file with birth history file. Here, only those births were taken into account which has taken place between inter-survey periods. Thus, after all, filtering the sample size reduces to 7,166 children out of them 3,905 belong to under age five with the information in all variable of interest including anthropometric measures.

### Outcome variables

In the present paper, child healthcare and growth outcomes are taken as child immunization, exclusive breastfeeding and nutritional status (figure 1). The child immunization was calculated using four vaccines BCG, DPT (1 to 3), Polio (1 to 3) and measles for children aged 0-5 years. Only those children were considered fully immunized who received all four vaccines (12). In case of breastfeeding only those children were considered exclusively breastfed who were breastfeeding for more than six months without any supplementary food (13). The nutritional status of children under age five years is calculated using STATA-macro file provided by (14). This STATA macro file calculates the height for age z-scores and weight for age z-scores for the children.

**Figure 1:**
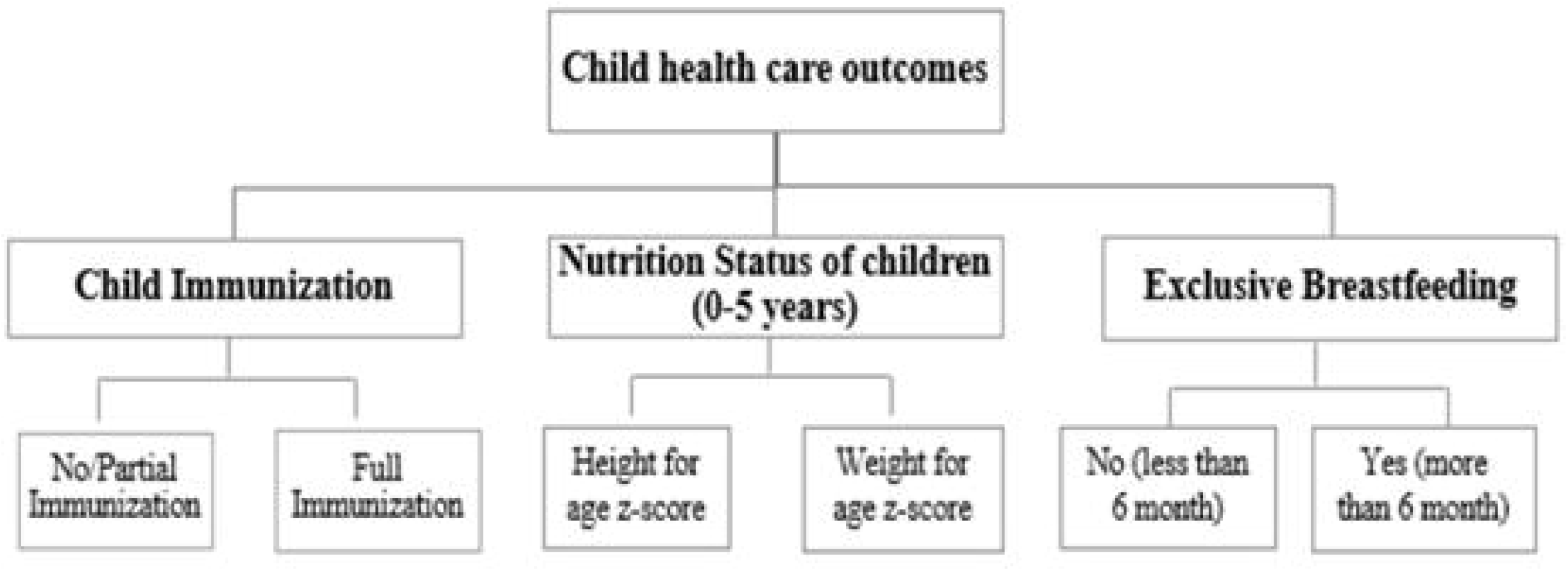
Child health care and growth-related outcomes

### Measurement of childbearing intention

All births to women who reported that they did not want to have a/additional child in the first round (IHDS-I) but gave birth during the inter-survey period, were classified as unintended births. If the woman wanted to have a/additional child in the first round, she was further asked how many did she want? These children are referred as the desired number of additional children and were matched with their actual number of children born during the inter-survey period. The number of births exceeding the desired number of children reported by women was classified in unintended births. Remaining births were classified as intended births

### Other independent variables

The socioeconomic and demographic variables in this study include, number of children survived (0-2, 2-4, 4+), sex of the child (Boy, Girl), Religion (Hindu, Muslim, Others), Caste groups (Forward caste/General, Other Backward caste, Scheduled caste, Schedule Tribe, No caste), Mother’s education (No education, Primary, Secondary, Higher), Place of residence (Rural, Urban), Empowered Action Group states (Non-EAG, EAG) and Wealth Index (low, medium, high).

### Statistical analysis

We employed bivariate with chi-square test and multivariate analyses to accomplish the study objectives. A multivariate logistic regression model was applied to analyse the effect of unintended and intended childbearing on child-immunization and exclusive breastfeeding after controlling other socioeconomic and demographic variables. For this purpose, both outcome variables were classified in binary form as-fully immunized (1-yes, 0-no) and exclusively breastfed (1-yes, 0-no). Further, to assess the effect of childbearing intention on child nutritional status, linear regressions were applied using height-for-age-z-score (HAZ) and weight-for-age-z-score (WAZ) as outcome variables and other socioeconomic and demographic variable as independent variables. The whole analysis was performed using STATA 13.

## RESULTS

Nearly 68% of intended births and 57% of unintended births were fully immunized (Table 1). These differences were statistically significant (p-value <0.001). Moreover, around 32.9% of intended births in contrast to 29.9% of unintended births were exclusively breastfed (p<0.05). Further, a higher mean value of HAZ (−1.75) among intended births as compared to (−2.11) among unintended births. Mean z score for weight for age (WAZ) was significantly lower among unintended births (−1.49) as compared to intended births (−1.14).

**Table 1:**
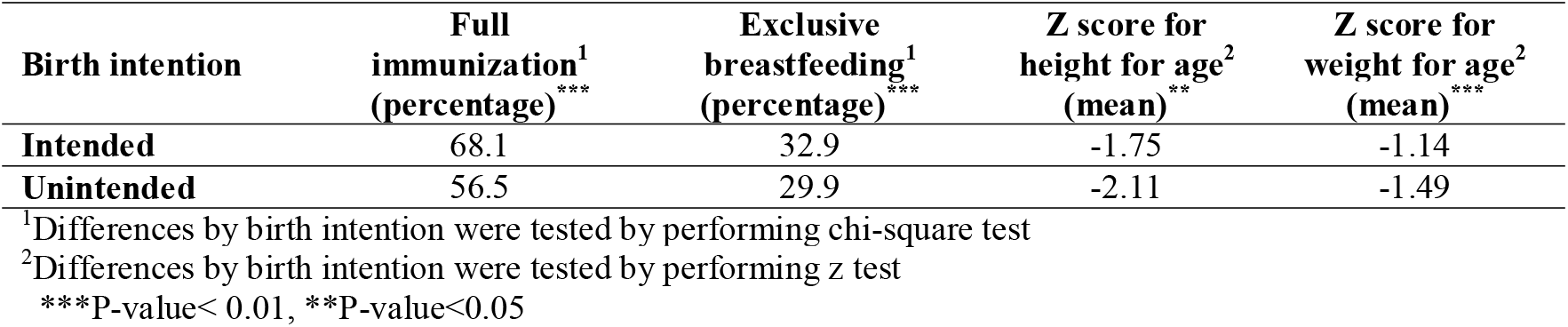
Child health care outcomes by birth intention reported by mothers

The result reveals that the odds of child immunization and exclusive breastfeeding reduced by 0.165 (p<0.001) and 0.109 (p<0.05) times respectively if the child was unintended (Table 2). Mother having 4 and above children survived were less likely to receive full immunization compared to others. Similarly, children from Muslim community were less likely to receive full immunization and exclusive breastfeeding compared to other communities. However, the children of educated mothers with better wealth condition were more likely to receive full immunization than their counterparts. Moreover, the odds of exclusive breastfeeding were 0.291 (p<0.001) times lower among Empowered Action Group (EAG) states corresponding to those who were living in Non-EAG states. As far as caste of the children is concerned, children who belonged to scheduled tribe and scheduled caste were less likely to receive full immunization as compared to children from forward/general caste.

**Table-2:** Odds ratios (OR with 95% confidence intervals) from multivariate regression analyses examining associations between selected characteristics and child immunization and exclusive breastfeeding

| **Covariates** | **Child immunization (C.I.)** | **Exclusively breastfed (C.I.)** |
| --- | --- | --- |
| **Birth Intention** |  |  |
| Intended<sup>(R)</sup> |  |  |
| Unintended | 0.835*** (0.739, 0.931) | 0.891** (0.792, 0.990) |
| **Children Survival** |  |  |
| 1-2<sup>(R)</sup> |  |  |
| 2-4 | 1.005 (0.898, 1.240) | 1.141 (0.973, 1.309) |
| 4+ | 0.693*** (0.563, 0.824) | 1.111 (0.904, 1.319) |
| **Sex of the child** |  |  |
| Male<sup>(R)</sup> |  |  |
| Female | 1.02 (0.908, 1.132) | 0.951 (0.848, 1.054) |
| **Caste Groups** |  |  |
| General<sup>(R)</sup> |  |  |
| Other Backward Caste (OBC) | 1.003 (0.849, 1.158) | 1.096 (0.938, 1.254) |
| Scheduled Caste (SC) | 0.775** (0.637, 0.914) | 0.941 (0.783, 1.100) |
| Scheduled Tribe (ST) | 0.723** (0.567, 0.879) | 1.135 (0.895, 1.375) |
| No Caste | 0.823 (0.387, 1.259) | 1.756 (0.890, 2.623) |
| **Religion** |  |  |
| Hindu<sup>(R)</sup> |  |  |
| Muslims | 0.648*** (0.549, 0.747) | 0.658*** (0.556, 0.761) |
| Others | 0.973 (0.654, 1.292) | 0.521*** (0.367, 0.675) |
| **Mother's Education** |  |  |
| No Education<sup>(R)</sup> |  |  |
| Primary | 1.837*** (1.551, 2.123) | 1.107 (0.933, 1.282) |
| Secondary | 2.100*** (1.771, 2.429) | 1.122 (0.952, 1.293) |
| Higher | 3.557*** (2.521, 4.593) | 1.207 (0.926, 1.489) |
| **EAG states** |  |  |
| Non-EAG<sup>(R)</sup> |  |  |
| EAG | 0.374*** (0.282, 0.466) | 0.709*** (0.574, 0.844) |
| **Place of Residence** |  |  |
| Rural<sup>(R)</sup> |  |  |
| Urban | 0.673 (0.232, 1.114) | 0.988 (0.854, 1.122) |
| **Wealth Index** |  |  |
| Low<sup>(R)</sup> |  |  |
| Medium | 1.277** (1.108, 1.446) | 1.173** (1.012, 1.335) |
| High | 1.386*** (1.164, 1.609) | 1.103 (0.921, 1.286) |
| **Constant** | 2.824 (2.136, 3.513) | 0.567 (0.433, 0.701) |

®= reference category and \*\*\*(p<0.001), \*\*(p<0.05), \*(p<0.10)

Table 3 portrays the determinants of child malnutrition. After controlling other factors, unintended children had significantly lower HAZ (−0.193, p<0.05) and WAZ (−0.172, p<0.05). However, scores were increased with mother’s education and households’ wealth index. Children who belonged to scheduled caste (−0.205, p<0.05) and scheduled tribes (−0.267, p<0.05) had lower HAZ as compared to children from general caste.

**Table-3:** Association between birth intention and z-score for height for age (HAZ) and weight for age (WAZ): Results from linear regression (Beta coefficients)

| Covariates | HAZ | WAZ |
| --- | --- | --- |
| **Birth Intention** |  |  |
| Intended<sup>(R)</sup> |  |  |
| Unintended | -0.193** (-0.308, -0.077) | -0.172** (-0.335, -0.010) |
| **Children Survival** |  |  |
| 1-2<sup>(R)</sup> |  |  |
| 2-4 | -0.065 (-0.220, 0.088) | -0.011 (-0.229, 0.206) |
| 4+ | -0.105 (-0.291, 0.080) | -0.003 (-0.265, 0.259) |
| **Sex of the child** |  |  |
| Male<sup>(R)</sup> |  |  |
| Female | -0.045 (-0.138, 0.048) | -0.052 (-0.155, 0.051) |
| **Caste Groups** |  |  |
| General<sup>(R)</sup> |  |  |
| Other Backward Caste (OBC) | -0.157** (-0.301, -0.013) | 0.047 (-0.155, 0.250) |
| Scheduled Caste (SC) | -0.205** (-0.371, -0.038) | -0.130 (-0.365, 0.104) |
| Scheduled Tribe (ST) | -0.267** (-0.482, -0.052) | 0.110 (-0.193, 0.413) |
| No Caste | -0.814** (-1.467, -0.160) | -0.667 (-1.696, 0.362) |
| **Religion** |  |  |
| Hindu<sup>(R)</sup> |  |  |
| Muslims | 0.077 (-0.068, 0.224) | -0.022 (-0.229, 0.184) |
| Others | 0.536*** (0.238, 0.834) | 0.333 (-0.089, 0.756) |
| **Mother's Education** |  |  |
| No Education<sup>(R)</sup> |  |  |
| Primary | 0.137* (-0.019, 0.294) | 0.163 (-0.057, 0.384) |
| Secondary | 0.201** (0.049, 0.353) | 0.183* (-0.030, 0.398) |
| Higher | 0.360** (0.115, 0.606) | 0.435** (0.090, 0.780) |
| **Place of Residence** |  |  |
| Rural<sup>(R)</sup> |  |  |
| Urban | 0.114* (-0.020, 0.250) | -0.045 (-0.235, 0.144) |
| **EAG states** |  |  |
| Non-EAG<sup>(R)</sup> |  |  |
| EAG | -0.025 (-0.220, 0.170) | -0.117 (-0.338, 0.103) |
| **Wealth Index** |  |  |
| Low<sup>(R)</sup> |  |  |
| Medium | 0.108 (-0.021, 0.237) | 0.108 (-0.072, 0.290) |
| High | 0.158* (-0.004, 0.321) | 0.285*** (0.055, 0.515) |
| **Constant** | -1.587 (-1.830, -1.344) | -1.911 (-2.256, -1.566) |

®= reference category and \*\*\*(p<0.001), \*\*(p<0.05), \*(p<0.10)

## DISCUSSION

The present study sheds light on adverse health outcomes of children born as unintended births using prospective approach. India, a country which has the highest number of underweight and anemic children, the presence of unintended birth will more likely to worsen the child physical and psychological development. Singh, Chalasani (9) in their study mentioned that each year there are estimated 26 million births in India, if one fifth of all births are unintended every year then at least some of them will suffer from adverse health outcomes, which is also likely to be seen in our study that unintended children are less likely to exclusively breastfed and received full immunization. The present finding is similar to previous studies which suggests that unintended pregnancy is associated with late initiation of prenatal care and inadequate childhood vaccination (4, 5, 15-17). The present analysis vividly brings out the fact that children who were reported as unintended are more likely to be stunted and underweight. While previous studies only reported that unintended births are significantly more likely to be stunted than intended (8, 9, 18). This work has examined the association between unintended births assessed prospectively and poor child growth using the longitudinal dataset for all India.

The tendency of low exclusive breastfeeding and full immunization among unintended children reflects that women are not giving proper care to their children even from the early stages of their life. One reason could be the less emotional attachment from mother to their children as they are unintended. In the present study the likelihoods of full immunisation, exclusive breastfeeding, weight for age z-score and hight for age z-score are not significant for female children. But, the maltreatment of female children has been very common phenomenon throughout the country as it is believed that female children are burden to the family. This is also the reason for low sex ratio, and child ratio in India. Further, it is unavoidable truth that female foeticide and under-nourished female children are quite prevalent, in major parts of our country(18-20). Moreover, above result shows that the likelihood of children’s health reduces with more number children, which simply reflects that mother could not be able to take care of their next kin if she is already having 2 or more children. Further, taking care of more than 2 children is very difficult socially and economically. Therefore, with growing living cost and education cost, people in India are becoming more sceptical for intending another child, resulting in comparatively low total fertility rate (TFR) in India. The recent report of national family and health survey (NFHS-4) shows that the current TFR of India is 1.8 which has reduced from 2.1 in NFHS-3. Therefore, it is quite likely that unintended children belong to rural area and low-economic background are more likely to have poor health conditions.

Further, the current study has various limitation like not including other child development indicators viz. education and their cognitive skills. Moreover, the seven years’ duration of the survey is a longer duration when woman’s childbearing intention would have changed and thus, child healthcare and growth would not be so influenced by her previously stated intention to have children. Further, father’s intention would also have influenced child well-being.

Apart from these limitations, the study has provided an insight that unintended pregnancy is serious issue for a country like India as it is related with adverse maternal and child health outcomes. Therefore, the present study contributes in body of evidence supporting the fact the unintended pregnancy is not only concerned with fertility levels but also crucial for public health challenges.

## CONCLUSION

This study provides clear evidence that the children born as unintended face series health implications. As children are unintended, most of the mothers do not pay attention to them and due to this behaviour, they receive inadequate immunization and exclusive breastfeed during the early childhood development period. The same can be seen in children’s physical development, as unintended births are more likely to be stunted and underweight. The poor physical development of children may hinder their cognitive ability, which also results in poor academic performance, and school dropouts (21-25). In many occasions, unintended births result in neo-natal mortality and infant mortality in India (9, 26-28). Therefore, averting unwanted births at individual level enhances the well-being of women and children. There should be renewed and revitalized attempts to help couples especially women to achieve their reproductive goals and avoid unwanted pregnancy.

### Contributors

All authors Poulomi Chowdhury, Mausam Kumar Garg and Md Illias Kanchan Sk, contributed to conceptualizing, writing and editing this paper. Data analysis was performed by Poulomi Chowdhury and Mausam Kumar Garg.

## Data Availability

The data was downloaded from IHDS website. This data is open access and available for anyone to conduct research.

https://ihds.umd.edu/data/data-download

## ETHICS APPROVAL

This research paper is based on secondary data which is available in the public domain for research use, and therefore there is no formal approval from the institutional review board is required.

## ACKNOWLEDGEMENT

This research is the part of M.Phil. Dissertation conducted within the International Institute for Population Sciences. I thank my supervisor Dr. Preeti Dhillon of the Department of Mathematical Demography for their continuous insight and guidance during my research.

## DATA SHARING STATEMENT

No additional data are available.

## REFERENCES

1. Gipson JD, Koenig MA, Hindin MJ. The effects of unintended pregnancy on infant, child, and parental health: a review of the literature. Studies in family planning. 2008;39(1):18–38.

2. Sedgh G, Singh S, Hussain R. Intended and unintended pregnancies worldwide in 2012 and recent trends. Studies in family planning. 2014;45(3):301–14.

3. World Health Organisation. Preventing Unsafe Abortion 2019 [Available from: https://www.who.int/news-room/fact-sheets/detail/preventing-unsafe-abortion.

4. Logan C, Holcombe E, Manlove J, Ryan S. The consequences of unintended childbearing. Washington, DC: Child Trends and National Campaign to Prevent Teen Pregnancy. 2007;28:142–51.

5. Singh A, Singh A, Thapa S. Adverse consequences of unintended pregnancy for maternal and child health in Nepal. Asia Pacific Journal of Public Health. 2015;27(2):NP1481–NP91.

6. Lindberg L, Maddow-Zimet I, Kost K, Lincoln A. Pregnancy intentions and maternal and child health: an analysis of longitudinal data in Oklahoma. Maternal and child health journal. 2015;19(5):1087–96.

7. Hall JA, Benton L, Copas A, Stephenson J. Pregnancy intention and pregnancy outcome: systematic review and meta-analysis. Maternal and child health journal. 2017;21(3):670–704.

8. Singh A, Upadhyay AK, Singh A, Kumar K. The association between unintended births and poor child development in India: Evidence from a longitudinal study. Studies in family planning. 2017;48(1):55–71.

9. Singh A, Chalasani S, Koenig MA, Mahapatra B. The consequences of unintended births for maternal and child health in India. Population Studies. 2012;66(3):223–39.

10. Singh A, Singh A, Mahapatra B. The consequences of unintended pregnancy for maternal and child health in rural India: evidence from prospective data. Maternal and child health journal. 2013;17(3):493–500.

11. Indian Human Development Survey. 2011 [updated 09 August 2018. Available from: https://www.icpsr.umich.edu/icpsrweb/DSDR/studies/34480.

12. World Health Organisation. WHO vaccine-preventable diseases: monitoring system: 2009 global summary. World Health Organization; 2009.

13. santé Omdl, Staff WHO, Organization WH, UNICEF., UNAIDS. Global strategy for infant and young child feeding: World Health Organization; 2003.

14. World Health Organisation. Child Growth Standards 2007 [Available from: https://www.who.int/childgrowth/software/en/.

15. Delgado-Rodriguez M, Gómez-Olmedo M, Bueno-Cavanillas A, Gálvez-Vargas R. Unplanned pregnancy as a major determinant in inadequate use of prenatal care. Preventive medicine. 1997;26(6):834–8.

16. Kost K, Landry DJ, Darroch JE. Predicting maternal behaviors during pregnancy: does intention status matter? Family planning perspectives. 1998:79–88.

17. Marston C, Cleland J. Do unintended pregnancies carried to term lead to adverse outcomes for mother and child? An assessment in five developing countries. Population studies. 2003;57(1):77–93.

18. Doskoch P. Unplanned pregnancy linked to poor child health in India. International Perspectives on Sexual and Reproductive Health. 2012;38(4):223.

19. Jena KCJOR, December Issue. Female foeticide in India: A serious challenge for the society. 2008:8–17.

20. Mishra V, Roy TK, Retherford RDJP, review d. Sex differentials in childhood feeding, health care, and nutritional status in India. 2004;30(2):269–95.

21. Gashu D, Stoecker BJ, Bougma K, Adish A, Haki GD, Marquis GS. Stunting, selenium deficiency and anemia are associated with poor cognitive performance in preschool children from rural Ethiopia. Nutrition journal. 2015;15(1):38.

22. Crookston BT, Dearden KA, Alder SC, Porucznik CA, Stanford JB, Merrill RM, et al. Impact of early and concurrent stunting on cognition. Maternal & Child Nutrition. 2011;7(4):397–409.

23. Black RE, Allen LH, Bhutta ZA, Caulfield LE, De Onis M, Ezzati M, et al. Maternal and child undernutrition: global and regional exposures and health consequences. The lancet. 2008;371(9608):243–60.

24. Fernald LC, Kariger P, Hidrobo M, Gertler PJ. Socioeconomic gradients in child development in very young children: Evidence from India, Indonesia, Peru, and Senegal. Proceedings of the National Academy of Sciences. 2012;109(Supplement 2):17273–80.

25. Connell M. Evaluating the Effects of Nutritional Intake During Adolescence on Educational Attainment and Labor Market Earnings as an Adult. 2018.

26. Lawn JE C. LancetNeona talSurvivalSteeringTeam. 4millionneonataldeat hs: when? Where? Why. Lancet. 2005:5–11.

27. Joyce TJ, Kaestner R, Korenman S. The effect of pregnancy intention on child development. Demography. 2000;37(1):83–94.

28. Chalasani S, Casterline JB, Koenig MA, editors. Consequences of unwanted childbearing: a study of child outcomes in Bangladesh. annual meeting of the Population Association of America, New York; 2007.

